# HIV prevalence and associated factors among married women, Mozambique, 2015

**DOI:** 10.1101/2023.06.01.23290844

**Authors:** Samuel Nuvunga, Denise Chitsondzo Langa, Cynthia Semá Baltazar, Jahit Sacarlal, Erika Rossetto

## Abstract

**Background:** The 2021 human immunodeficiency virus and acquired immunodeficiency syndrome (HIV/AIDS) global report indicated that women and girls in sub-Saharan Africa remained the most affected groups, accounting for 62% of new infections in the region. In 2021 in Mozambique, the HIV prevalence rate among the adult population was 12.5%, with a rate of 15.4% among women of reproductive age. Our study assessed HIV prevalence and associated factors among married women in Mozambique.

**Methodology:** A cross-sectional epidemiological study was carried out using secondary data extracted from the 2015 Immunization, Malaria and HIV/AIDS (IMASIDA) survey in Mozambique. Married women (civil marriage and common law marriage) of reproductive age (15-49 years) were included. We applied sample weights for data analysis.

**Results:** Out of the 3,006 married women included in the study, 20.1% fell within the 20-24 age group (n=603), and the average age was 30 years (SD=8.9 years). HIV prevalence was 6.9%. Factors significantly associated with HIV infection were age group 35-49 years (aOR=2.5; CI [1.3-4.6]; p=0.005) compared to age group 15-24 years, lack of formal education and primary education compared to higher education (aOR 7.7; CI [1.1-52.9], p=0.038 and aOR=9.8; CI [1.6-60.1]; p=0.014 respectively), having an uncircumcised partner (aOR=1.9; CI [1.2-3.1]; p=0.008), and having three or more lifetime sex partners compared to one sex partner (aOR=3.6; CI [2.9-7.3]; p< 0.001). Women who were in one lifelong union had a lower risk of HIV positivity compared to women who hadmore than one union (aOR=0.5; CI[0.3-0.8] p=0.005).

**Conclusion:** The factors that contributed to a greater odds of HIV-positivity in this group of married Mozambican women were older age, low levels of education, having an uncircumcised partner and having more than one sexual partner throughout one’s lifetime. In order to have maximum effect, HIV prevention and control campaigns in Mozambique should be tailored tothese “higher-risk” populations.

## Background

Human immunodeficiency virus (HIV) remains a worldwide public health problem and has implications for socio-economic development, especially in developing countries. According to the 2022 UNAIDS data, about 38.4 million people worldwide were living with HIV, with 1.5 million new HIV infections in 2021[1].

A report by the Joint United Nations Programme on HIV/AIDS (UNAIDS) in 2020 revealed that inequalities between men and women put women at greater risk of exposure to HIV/AIDS. Despite representing only 10% of the Sub-Saharan Africa population, adolescents and young women aged 15-24 alone accounted for 25% of new HIV infections in 2019 [2]. From 2018 to 2021, women of reproductive age accounted for a disproportionate number of infections, with a prevalence three times higher than men in East and Southern Africa and South America [3,4,5]. Women in sub-Saharan Africa remain the most affected by HIV, accounting for 59% of all new infections in the region in 2019 [6]. Gender discrimination and violence, gaps in education, lack of economic autonomy and lack of protection of sexual health rights are leading to an increased risk of HIV exposure for women [2,7].

HIV prevalence among Mozambican adults was estimated at 10.6% in 2015, which places Mozambique among the countries most affected by the AIDS epidemic[8]. The HIV epidemic in Mozambique is not evenly distributed; adolescents and young adults are among the most vulnerable groups. In 2015, the estimated prevalence of HIV among adolescents and young adults aged 15 to 24 was 6.9% [9].

There are few studies related to HIV among married women in Mozambique, despite increases in the number of HIV infections within the population. Married women may underestimate their risk of contracting HIV if they presume that their partner is monogamous or they may find it difficult to negotiate safe sex practices with the partner due to fears of violence or separation[10]. This cross-sectional study aimed to estimate the prevalence of and analyze risk factors associated with HIV/AIDS among married women of reproductive age in Mozambique in 2015.

## Methods

### Study design

This study analyzed data from the second national and demographic survey of HIV and AIDS prevalence, risk behavior and information (IMASIDA) carried out in 2015[9]. the IMASIDA survey was cross-sectional and carried out in two phases with the application of stratification methods and cluster sampling to ensure that, for each province, inferences were possible with almost the same precision. Enumeration Areas (AEs), households and individuals were primary sampling units, secondary sampling units and tertiary sampling units, respectively[9].

The IMASIDA survey randomly selected 307 AEs from 45,000 AEs defined according to the 2007 General Population and Housing Census, 134 AEs from urban areas and 173 AEs from rural areas. A fixed number of households were systematically selected in each AE[9]. Men and women aged between 15 and 59 years participated in individual interviews and provided a blood sample for HIV testing. Participants who declared being HIV-positive were not tested for HIV.

During individual interviews, respondents were asked whether they were married and, if so, who their spouse was. If the spouse’s name was mentioned in the household questionnaire, the interviewer noted the spouse’s residential line number on the individual questionnaire. Through a methodology standardized by the Demographic and Health Surveys (DHS Program), cohabiting couples were matched, allowing for an analysis of their serology and other characteristics. The dataset for this survey can be accessed at https://dhsprogram.com/data/.

### Sample strategy

The primary database of IMASIDA was composed of 7,749 women aged between 15 and 59 years old, of which 2,823 were married. we restricted our population sample to include only married women of reproductive age (15-49 years). The final unweighted sample consisted of 2,669 women. and the final weighted sample was calculated as 3,006 women.

Due to the unproportionate assignment of samples by districts and differentials in response rates, the sampling weighting of the results of IMASIDA 2015 should be used in all analyses to ensure nationally representative results. For our study, the data were calculated using the same weights applied to the IMASIDA results. For more details about sample probabilities and sampling weights visit the IMASIDA report.

### Data analysis

Data were analyzed using STATA software version 16.1. Descriptive analyses were conducted to summarize socio-demographic and behavioral characteristics using frequencies, percentages, averages, and standard deviations. Pearson’s chi-square tests were used in bivariate analyses to verify the association between categorical variables.

In a first phase, bivariate analysis of each factor in association with HIV was performed. Factors that presented evidence of association (p<0.20) with positive HIV test results in the bivariate analysis were considered a hypothesis of association and included in the multivariate analysis to obtain the association of interest and in the end a multivariate logistic regression was used to estimate adjusted odds ratios. A 95% confidence interval and a significance level of p < 0.05 were used to define statistical significance in the final model, noting that all analyzes were adjusted to take into account the complex sample design.

### Ethical considerations

The final databases of the DHS Program surveys are public on the website (www.dhsprogram.com) and do not violate the principle of ethics and confidentiality because the data are anonymous and without any other potential identifiers of the participants. IMASIDA, 2015 was implemented according to Mozambican and international ethical standards and had the approval of the Mozambique National Bioethics Commission for Health (CNBS), code IRB0002657, with reference 262/CNBS/14. IMASIDA participants gave free and informed consent prior to participation.

As this is a secondary study, the database is provided by the National Institute of Health (NIH) to researchers interested in secondary studies is anonymous and not relevant and has no participant identifiers.

## Results

### Sociodemographic characteristics

Our study consisted of 3,006 married women, where 59% (1,777/3,006) were officially married and 41% (1,229/3,006) were in common-law marriages. Mean age was 30 years (STD = 8.9). Thirty five percent (1060/3,006) were between 15 and 24 years old. About 85% (2,546/3,006) spoke a local tribal language as their native language compared to 6% (175/3,006) who spoke Portuguese as their native language.

Most women were Catholic (32%; 975/3,006), followed by Protestant (16%; 478/3,006). The study found that 55% of women had an elementary education (1660/3,006) and that 32% of women had no formal education (946/3,006). Forty-six percent (1384/3,006) of women fell within the bottom two wealth quintiles (Table 1).

**Table I.** Sociodemographic characteristics among married women in reproductive age, Mozambique-2015.

| Characteristics | * Civil marriage | † Common-law marriage | Total |
| --- | --- | --- | --- |
|  | (n = 1777) | (n = 1229) | (N = 3006) |
|  | n (%) | n (%) | n (%) |
| **Age group (years)** |  |  |  |
| 15-24 | 615 (34.6) | 445 (37.0) | 1060 (35.3) |
| 25-34 | 558 (31.4) | 420 (34.2) | 978 (32.5) |
| 35-49 | 604 (33.9) | 374 (30.4) | 978 (32.5) |
| **Main language spoken** |  |  |  |
| Portuguese | 84 (4.7) | 91 (7.4) | 175 (5.8) |
| Mother tongue(native language)++ | 1528 (86.0) | 1018 (82.8) | 2546 (84.7) |
| Other | 165 (9.3) | 130 (10.6) | 295 (9.8) |
| **Religion** |  |  |  |
| Catholic | 668 (37.6) | 307 (24.9) | 975 (32.4) |
| Protestant | 249 (14.0) | 232 (17.9) | 481 (16.0) |
| Evangelical | 112 (6.3) | 188 (15.3) | 300 (10.0) |
| Zione | 100 (5.6) | 203 (16.5) | 303 (10.1) |
| Other | 534 (30.1) | 223 (18.1) | 757(25.2) |
| None | 114 (6.4) | 86 (7.0) | 200 (6.6) |
| **Education level** |  |  |  |
| No education | 618 (34.8) | 328 (26.7) | 946 (31.5) |
| Primary | 944 (53.1) | 716 (58.3) | 1660 (55.2) |
| Secondary | 194 (10.9) | 185 (15.1) | 379 (12.6) |
| Higher | 21 (1.2) | 10 (0.8) | 31 (1.0) |
| **Employment** |  |  |  |
| No | 1164 (65.5) | 684 (55.6) | 1848 (61.5) |
| Yes | 613 (34.5) | 555 (45.2) | 1168 (38.9) |

| **Area of residence** |  |  |  |
| --- | --- | --- | --- |
| Urban | 441 (24.8) | 403 (32.8) | 844 (28.1) |
| Rural | 1336 (75.2) | 836 (68.0) | 2172 (72.3) |
| **Zone of residence** |  |  |  |
| North | 1031 (58.0) | 372 (30.3) | 1403 (46.7) |
| Center | 650 (36.6) | 411 (33.4) | 1061 (35.3) |
| South | 96 (5.4) | 456 (37.1) | 552 (18.4) |
| **Quintile of wealth** |  |  |  |
| Poorest | 451 (25.4) | 211 (17.1) | 662 (22.0) |
| Poorer | 498 (28.0) | 224 (18.2) | 722 (24.0) |
| Middle | 373 (21.0) | 270 (21.9) | 643 (21.4) |
| Richer | 261 (14.7) | 258 (21.0) | 519 (17.3) |
| Richest | 194 (10.9) | 276 (22.5) | 470 (15.6) |

\* Is a legally recognized marriage between two people who have not purchased a marriage license or had their marriage solemnized by a ceremony
† Is a marriage performed, recorded and recognised by a government official. Such a marriage may be performed by a religious body and recognised by the state, or it may be entirely secular.
†† The term "mother tongue" refers to a person's native language — that is, a language learned from birth. Also called a first language, dominant language, home language, and native tongue (although these terms are not necessarily synonymous).

### Factors associated with HIV prevalence

Among married women of reproductive age (both those in officially and common law marriages), HIV prevalence was 6.9% (95% CI: 5.7-8.4). The lowest prevalence (0.1%) was among women with higher education and with one lifetime sexual partner. The highest prevalence (6.4%) was among women who reported having had an STI in the past 12 months (Table II).

**Table II.**
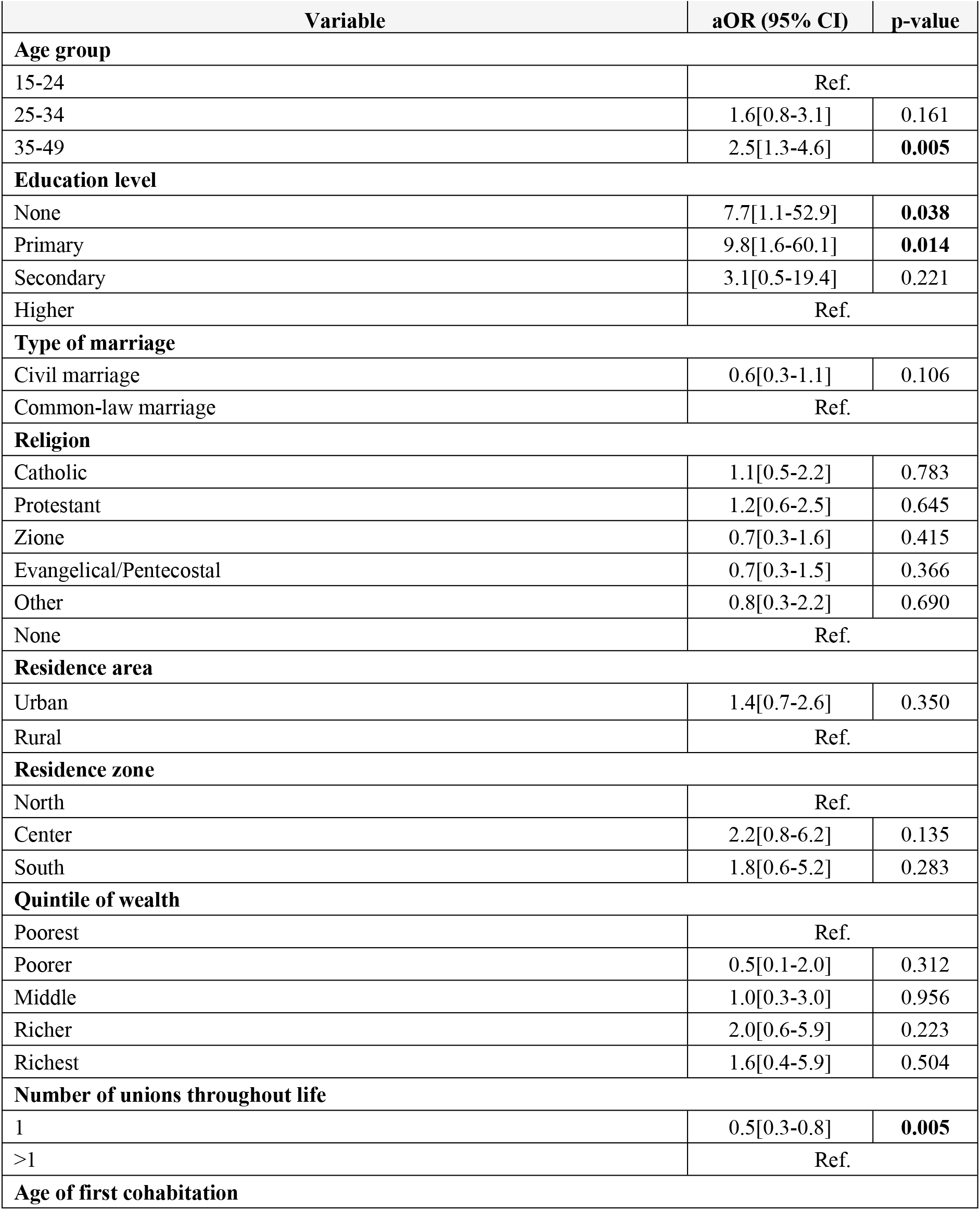
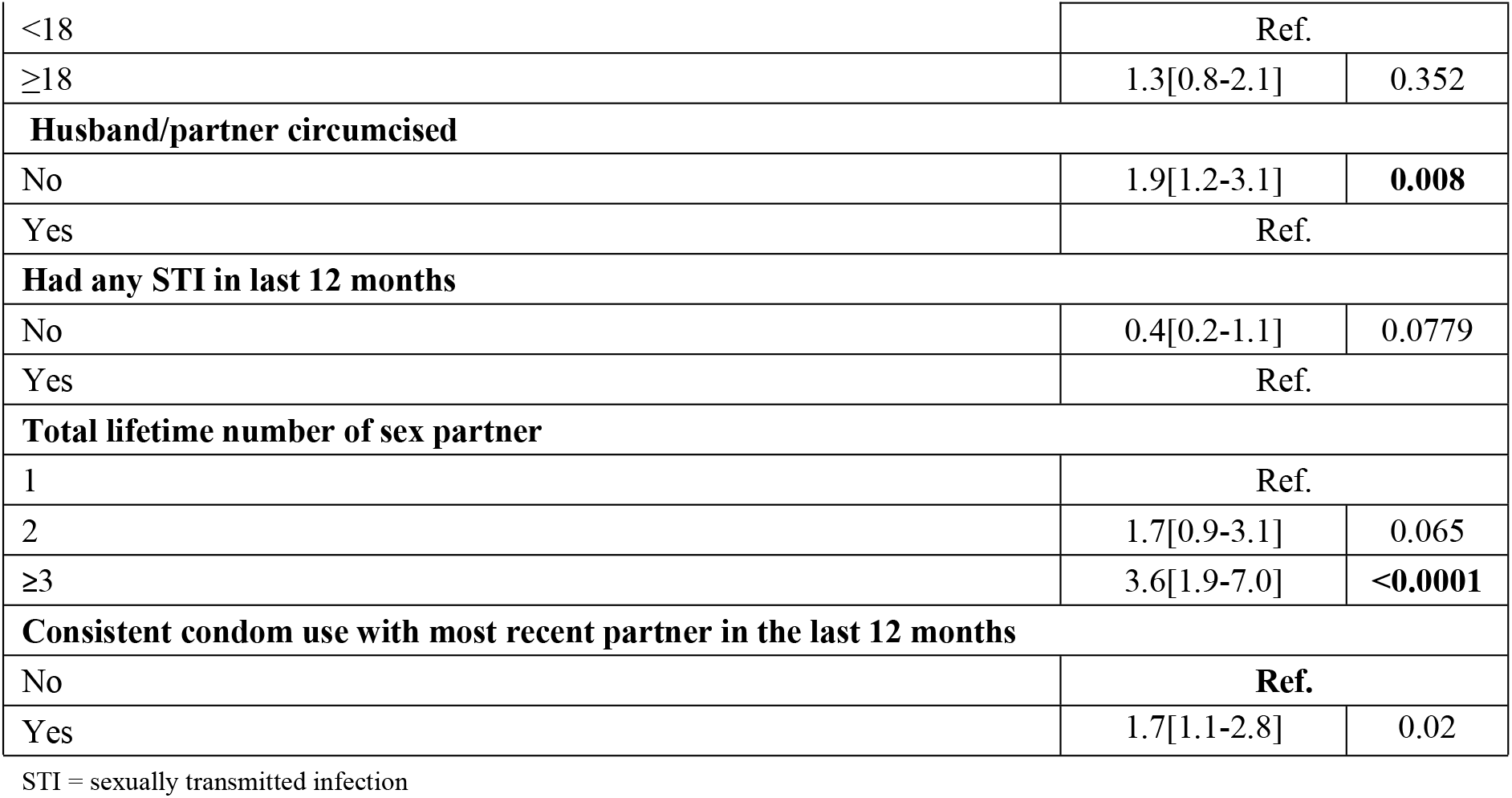
Multivariate analysis of factors associated with HIV infection among married women in reproductive age, Mozambique-2015. (n = 1,753)

Multivariate logistic regression analyses indicated that women aged 35 to 49 years were more likely to be infected with HIV compared to those aged 15 to 24 years (aOR=2.5[1.3-4.6], p=0.005).

The odds of being HIV-positive was significantly higher among women with no education (aOR=7.7[1.1-52.9], p=0.038) and among women with elementary education (aOR=9.8[1.6-60.1], p=0.014) versus those with higher education. The results also showed that women who reported having one union throughout their lives were 50% less likely to be HIV positive than those who reported more than one union throughout their lives (aOR=0.5[0.3-0.8], p=0.005).

Women who did not use condoms with their most recent partner in the past 12 months were more likely to be HIV-positive compared to those who used condoms (aOR=1.8[1.1-2.8], p<0.0001). Additionally, compared with women who reported having a circumcised partner, the odds of being HIV-positive were significantly higher in women who reported having an uncircumcised partner (aOR=1.9[1.2-3.1], p=0.008).

Women who reported more than three sexual partners in life were more likely to have an HIV-positive status compared to those who reported having only one sexual partner (aOR=3.6[1.9-7.0], p<0,0001).

Our findings showed that the odds of seropositivity were 1.7 times higher (p=0.02) in women who reported having used a condom consistently in their last relationship in the 12 months prior to the survey compared to who reported not using condoms (this odds increased to 2.9 in the bivariate model).Some sociodemographic and behavioral characteristics such as type of marriage, religion, area of residence, wealth quintile, age at first cohabitation and having had a sexually transmitted infection in the last 12 months were statistically significant in the bivariate model, however in the final model they did not show statistically significant association with HIV infection in the study population.

## Discussion

Our study results showed that married women contributed to the prevalence of HIV in Mozambique, although this prevalence was low (6.9%) compared to the prevalence of the general population at reproductive age at 15.4% [11,12].

In this study, HIV-positive serostatus was significantly associated with age; older married womenwere more likely to contract HIV than younger women, other studies have also [10,11,13], revealed an association between older age and greater likelihood of HIV, where the odds of being HIV-positive increase with increasing age. Some possible reasons for the higher HIV prevalence in older age groups are that these groups of women can be characterized as being at the peak of their sexual activity and have accumulated more time of exposure to HIV infection.

Lack or low levels of formal education were associated with an increased odds of being HIV-positive in this study. These findings are corroborated by studies conducted by Patra (2016), Ekholuenetale et al (2020) and Mocumbi et al (2017), which also show an association between low education levels and HIV risk [14–16]. This association may be related to the fact that individuals with less formal education may be less knowledgeable about how diseases are spread, prevented and controlled[17–21].

Married women who reported having been married more than once throughout their lives were more likely to be HIV positive. This may be related to the fact that they are more exposed to infection each time they change sexual partners[22,23]. A stable relationship often translates into consummating amarriage and, once there is trust and stability, there will be difficulties in using condoms. This can be motivated by trust in each new partner’s relationship. Having unprotected sex without prior testing for sexually transmitted diseases can thereby increase a woman’s risk of contracting HIV with each relationship[22,23].

It was not surprising that women who had an uncircumcised partner had a significantly higher odds of being HIV positive, given that male circumcision has been shown to provide a protective benefit against acquiring HIV [24–28]. However, we unexpectedly found that women who reported consistently using condoms were more likely to be HIV positive. We hypothesize that one reason for this finding could be that many women who consistently used condoms already knew their HIV-positive status, and therefore opted to regularly use condoms as a way of protecting their partner. Dias et al (2018) and Xavier’s (2020) studies on risk factors for HIV/AIDS in women who are married or in a stable partnership revealed that women who are married or in stable relationship feel inhibited from demanding that their partner use a condom, fearing that their partners may think that if they demand condom use it is because they are being unfaithful [11,22].

Expectations about adhering to traditional gender norms withinrelationships and difficulties within marital relationships can affect HIV infection susceptibility and the adoption of risk-free behaviors. It is crucially important to consider such marital partner dynamics when counseling women about HIV and designing HIV prevention and treatment programs.

Given that this study was based on nationally representative data, we believe that the results are generalizable to married women of reproductive age in Mozambique. However, the use of secondary data in this study limited the authors’ ability to control for biases in the data collection process and examine additional HIV risk factor variables that were not collected in the IMASIDA survey.

## Conclusion

Married women or in stable relationship contribute to the prevalence of HIV in Mozambique. The factors such as age, educational level, number of marriages throughout life, husband/partner circumcised and total lifetime number of sex partners were found to be significantly associated with HIV seropositivity among married women in Mozambique. These findings underscore the importance of developing HIV/AIDS prevention policies that consider individual, social, and cultural influences on the HIV epidemic and target higher-risk subpopulations of married women.

## Data Availability

All relevant data are within the manuscript and its Supporting Information files.

## Recommendations

It is recommended that HIV prevention and control programs focus their efforts on specific subgroups of the female married population, in order to achieve maximal impact.

## Acknowledgements

We are grateful for the Field Epidemiology Training Program (FETP) for monitoring the study, and for the willingness of INS to make the IMASIDA database available for analysis. Thank you to Hilenio Sabão for the explanation of how to manipulate IMASIDA data. We also thank the Zandile Nukeri for her support in data analysis and Paola Rullán Oliver, Arianna Unger and Peter Young for their review of the study analysis methods and English language.

## What is known about this topic

- Mozambique is one of the countries in the world with the highest HIV/AIDS prevalence rates among the population of reproductive age (15.4%)
- Married women of reproductive age contribute significantly to the prevalence of HIV/AIDS in Mozambique

## What the study adds

- The study demonstrates the importance of strengthening HIV/AIDS surveillance and health education activities among married women of reproductive age
- Some sociodemographic and behavioral characteristcs among married women of reproductive age were found to be strongly associated with an HIV-positive status

## Financing

This study was made possible due to the financial support for FELTP coordinated by the National Institute of Health of Mozambique through the Cooperative Agreement PEPFAR 5U2GGH000080.

## Authors’ contributions

SN conducted the study analysis and wrote the manuscript; DCL, PM and JS monitored the analysis, CSBand ER reviewed the manuscript.

